# Effect of introducing ethambutol and integrating drugs into fixed-dose tablets on mortality in patients with tuberculosis

**DOI:** 10.1101/2023.04.18.23288764

**Authors:** Fredi Alexander Diaz-Quijano, Patricia Bartholomay, Kleydson B. Andrade, Daniele M. Pelissari, Denise Arakaki-Sanchez, Fernanda D. Costa, Rejane Sobrino Pinheiro

## Abstract

At the end of 2009, due to the increase in primary resistance to isoniazid, the Brazilian Ministry of Health established changes in the treatment regimen for tuberculosis. The changes included the addition of ethambutol as the fourth drug in the intensive treatment phase and the integration of the four drugs into fixed-dose combination tablets. The introduction of fixed combination doses also led to changes in the dosage of isoniazid and pyrazinamide in the intensive and maintenance phases.

**Objective:** To estimate the effect of changing the tuberculosis treatment regimen on all-cause mortality and, secondly, outcomes such as mortality due to tuberculosis, cure, and loss to follow-up.

**Methods:** We compared the cohorts of people diagnosed with tuberculosis from 2008 to 2013, aged ten years or older, who started treatment for tuberculosis in Brazil before and after the change in the regimen adopted in 2009 (n: 145528 vs. 161264). Data were extracted from the Notifiable Diseases (Sinan) and Mortality (SIM) information systems. The missing data were imputed, and the effects were estimated using multilevel logistic models, with the state as the aggregation cluster. A directed acyclic graph guided the selection of covariates.

**Results:** The current (modified) regimen was not associated with significant changes in all-cause mortality (Relative Risk [RR]: 1.01; 95% confidence interval [95%CI]: 0.98 – 1.04), or tuberculosis mortality (RR: 0.98; 95%CI: 0.95 – 1.02). For cure, when transfers and missing outcome data (MOD) were considered an absence of outcome, there were no differences between treatments. When they were assumed as cured or imputed, the cure was less frequent in the current treatment. When transfers and MOD were imputed or considered an absence of loss to follow-up, the latter was more frequent in the current treatment. There were no differences in loss to follow-up between treatments when transfers and MOD were interpreted as that outcome.

**Conclusion:** The implementation of the modified treatment regimen was not associated with increased mortality in tuberculosis patients. Although there was a lower record of cures and a higher frequency of loss to follow-up during the second period, the sensitivity analysis indicated that a reduction in transfers and unknown outcomes could explain these associations.

## INTRODUCTION

More than 10 million new cases of tuberculosis and 1.2 million deaths associated with this disease are reported annually.^1^ Thus, before the COVID-19 pandemic, tuberculosis was the leading cause of death from infectious diseases, surpassing HIV/AIDS and malaria.^2^ Brazil is included in the list of countries with a high burden of tuberculosis (TB) and TB-HIV co-infection, according to the World Health Organization (WHO).^3^ In 2020, 66,819 new cases of TB were reported (31.6 cases/100,000 inhabitants), of which 70.1% were cured.^4^ Problems with adherence to treatment and the emergence of resistant strains, among other factors, are challenges for controlling the disease.^5,6^

At the end of 2009, due to the increase in primary resistance to the drugs isoniazid (H) and rifampicin (R), the Brazilian Ministry of Health promoted changes in the basic treatment regimen for tuberculosis for adolescents and adults (10 years or older).^7^ The first change was the introduction of ethambutol as the fourth drug in the intensive treatment phase (first two months) of the basic regimen. The second change introduced the combination fixed-dose tablet of the four drugs (4 in 1) for the intensive phase of treatment, which also led to changes in the dosage of isoniazid and pyrazinamide in the intensive and maintenance phases.^7^

The introduction of a fourth drug was intended to increase the therapeutic success and avoid increased multidrug resistance. The expected advantages of the new drug presentation included greater patient comfort by reducing the number of pills; made it impossible to take drugs in isolation, which could reduce the selection of resistant strains; and simplified pharmaceutical management at all levels.^7^ This was ultimately expected to lead to a decrease in patient mortality.

An ecological study, based on an interrupted time series analysis, did not show an association between treatment regimens and the frequency of outcomes, such as loss to follow-up or cure. Nevertheless, deaths from pulmonary tuberculosis significantly increased during the period of the new regimen.^8^ Interrupted time series analyzes can help to evaluate interventions when confounders do not change or change slowly over time.^9^ However, the determinants of the tuberculosis burden have a wide variability in the country and several epidemiological and socioeconomic phenomena may have affected the disease prognosis in recent decades.^10,11^

For this reason, to estimate the effects of the new treatment regimen on tuberculosis outcomes, controlling for potential confounding associated with the characteristics of the patients is justified. Faced with the difficulty of carrying out a clinical trial, for ethical and feasibility reasons, we performed an observational study with individual data to estimate the effect of changing the tuberculosis treatment regimen on all-cause mortality. Secondly, we evaluated the association between the mentioned regimen and other outcomes including mortality due to tuberculosis, cure, and loss to follow-up.

## METHODOLOGY

### Study design and population

We compared the cohorts of patients with tuberculosis who started treatment in Brazil, before and after the change in the regimen adopted in 2009. The study population corresponded to patients diagnosed with tuberculosis and age 10 years or older. Changes in treatment regimen were not indicated for children under ten years old.

### Inclusion criteria

new cases of tuberculosis notified to the public health surveillance system and who started their first treatment for tuberculosis during any of the compared periods.

### Exclusion criteria

- Patients who started treatment in a transitional period. This is because the classification of the treatment regimen was based on the availability of the medicines from the Ministry to the states (including the Federal District). Therefore, this transition period was considered to ensure that the new treatment regimen, including changes in the intensive and maintenance phase, was incorporated in all health institutions of the corresponding state. For the intensive phase, the patient was considered in a transition period when the date of diagnosis occurred between 30 days before and 120 days after sending the new regimen to the corresponding state. In the maintenance phase, a patient was in a transition period when the date of diagnosis occurred between 150 days before and 60 days after sending the new regimen to the state.
- Cases in which, after notification, the diagnosis of tuberculosis was ruled out.
- Pregnant women. This is because several variables of pregnancy that could affect the prognosis, including gestation age and co-interventions, could not be controlled because they were not part of the systematically collected information.
- Patients with inconsistent information, such as date of death before the initial treatment date.

### Information sources

Data on sociodemographic variables, clinical and laboratory covariates, and outcomes were extracted from the Information System for Notifiable Diseases (Sinan). In the Sinan, the outcomes of TB treatment include the options “death due to tuberculosis” and “death due to other causes.” However, this system is not the gold standard for notifying deaths in Brazil. Thus, it was necessary to qualify the death information with the Mortality Information System (SIM).

Since the databases (Sinan and SIM) do not have a single identification variable, a link was performed using deterministic and probabilistic strategies at several stages from the identification fields: patient’s name, mother’s name, date of birth, sex, county of residence, and address. The Sinan database from 2007 to 2015 and the SIM database from 2007 to 2016 were linked. Preprocessing of name fields was performed, eliminating punctuation and symbols not belonging to the Latin alphabet, treatment pronouns, phonemes (de, da[s], do[s]), and other information not belonging to the names.

The deterministic approach was carried out by applying blocking keys composed of the phonetic code of fragments containing the person’s first, middle, and last name, date of birth, and sex. Pairs of records for the same person were those whose full names had a Levenshtein distance of less than 3 for the patient’s and mother’s names. The unpaired records were submitted to probabilistic linking routines, performed in eight blocking steps, using a combination of variables: phonetic codes of the first and last fragments of the individual’s name and mother’s name, year of birth, sex, and state. It started with the most specific strategy, containing all the variables and eliminating them in the following steps. All strategies included the state as a blocking variable. Unrelated pairs at each blocking step proceeded to manual review.

The deterministic routines were performed using the Structured Query Language (SQL) programming language with PostGres software, version 1.22.2, and the probabilistic linkage was performed with OpenRecLink software, version 3.1.182.4147.

### Treatment groups

We considered the treatment regimen recommended by the Ministry of Health until 2009 as the reference (previous), which was indicated for adolescents and adults. The regimen consisted of two months of rifampicin, isoniazid, and pyrazinamide, followed by four months of rifampicin and isoniazid (2RHZ/4RH), with rifampicin and isoniazid available in fixed combination doses (300/200 mg or 150/100 mg capsules).

The first changes were made in the intensive phase and included: 1) the addition of ethambutol; 2) a reduction in the dose of isoniazid (75 mg for every 150 mg of rifampicin) to match international recommendations; and 3) the incorporation of the fixed-dose combination tablet containing the four drugs (rifampicin 150 mg, isoniazid 75 mg, pyrazinamide 400 mg, and ethambutol 275 mg).^12^ A second change was incorporated in the maintenance drugs with the same dose adjustment for isoniazid starting to be available associated with rifampicin in tablets containing fixed combination doses (75/150 mg, respectively). Thus, the “current regimen” incorporated changes in both the intensive and maintenance phases of treatment.

### Outcomes

Death from any cause during the first year after starting treatment was considered the primary outcome. In addition, independent analyzes were performed to assess the effect of the treatment regimen on the following secondary outcomes:

- Natural death, which we defined by excluding external causes of death;
- Death attributed to tuberculosis;
- Lost to follow-up, as registered in the Sinan form;
- Cure;
- A first composite outcome (No. 1) defined by the occurrence of any of the following unsuccessful endpoints: death from any cause, lost to follow-up, or treatment failure;
- A second composite event (No. 2) defined by the occurrence of any of the following: death from any cause, lost to follow-up, treatment failure, or unknown final situation (i.e., missing outcome data [MOD]). In this case, transfer was considered as absence from the event.

For each of the secondary outcomes of cure and lost to follow-up, sensitivity analyzes were performed 1) considering transfer and MOD as absence of the outcome; 2) considering transfer and MOD as occurrence of the outcome; and 3) considering both transfer and MOD as similar types of missing and making their corresponding outcome imputation. For the first composite outcome, transfer and MOD were considered absence, but an estimation was also performed by imputing these two component endpoints.

### Covariates

Several factors associated with prognosis could change over time through demographic and epidemiological transitions, affecting the comparability of the treatment groups. These variables include age, race, education, place of residence, special populations (such as the prison population), economic conditions (income), comorbidities, and medical history, including HIV/AIDS, alcoholism, diabetes, and mental illness. The extrapulmonary clinical form was considered an indicator of disease severity.

Directly observed treatment would be regarded as a cointervention that could be determined by factors, such as place of residence or belonging to special populations. As this variable refers to a result that would be evaluated throughout the treatment, it does not precede exposure and would be discarded as a confounder. However, its possible determinants, including residence and population characteristics, were considered during the analysis planning.

### Analysis

A description of the population was made in each of the periods. The covariates that should be conditioned to estimate the effect of the treatment regimen were identified based on a conceptual model represented in a directed acyclic graph (DAG). In this DAG, the change in treatment regimen was time dependent. Over time, the determinants of tuberculosis outcomes were expected to change. These risk factors included variables known to be associated with unfavorable outcomes, such as alcohol abuse, age, and sex, among others.^13–16^ Thus, the time period was a common determinant of both the treatment and the risk factors. However, the period should not be considered as a covariate because it overlapped with the treatment being evaluated. Therefore, controlling for confounding would depend on adjusting for risk factors associated with the outcome.

Although outcomes, such as lost to follow-up and death, may have different mechanisms, we considered that the effect on one necessarily affects the other. Therefore, in this graph, the concept of “prognosis” is represented to refer to any of the analyzed outcomes.

Consequently, the DAG indicated that the effect estimates should be adjusted for the following set of variables: age, HIV/AIDS, sex, diabetes, vulnerable groups (prison population), residence (state), alcoholism, mental illness, and clinical form of the disease. This DAG exhibited three testable independent implications that were evaluated and had p-values greater than 0.6. Therefore, we considered that the DAG was consistent with the data.^17^

### Covariate trends, residual confounding, and sample restriction

We observed that several variables required for confounding control were strongly determined by time (see examples in Figure 2). For that reason, we considered that even using adjustment strategies, there was a high risk of residual confounding associated both with measurement errors of these covariates (as can happen with the HIV/AIDS classification) and with unmeasured features of the concepts they represent, such as illicit drug use and other determinants of tuberculosis virulence.^13^

**Figure 1.**
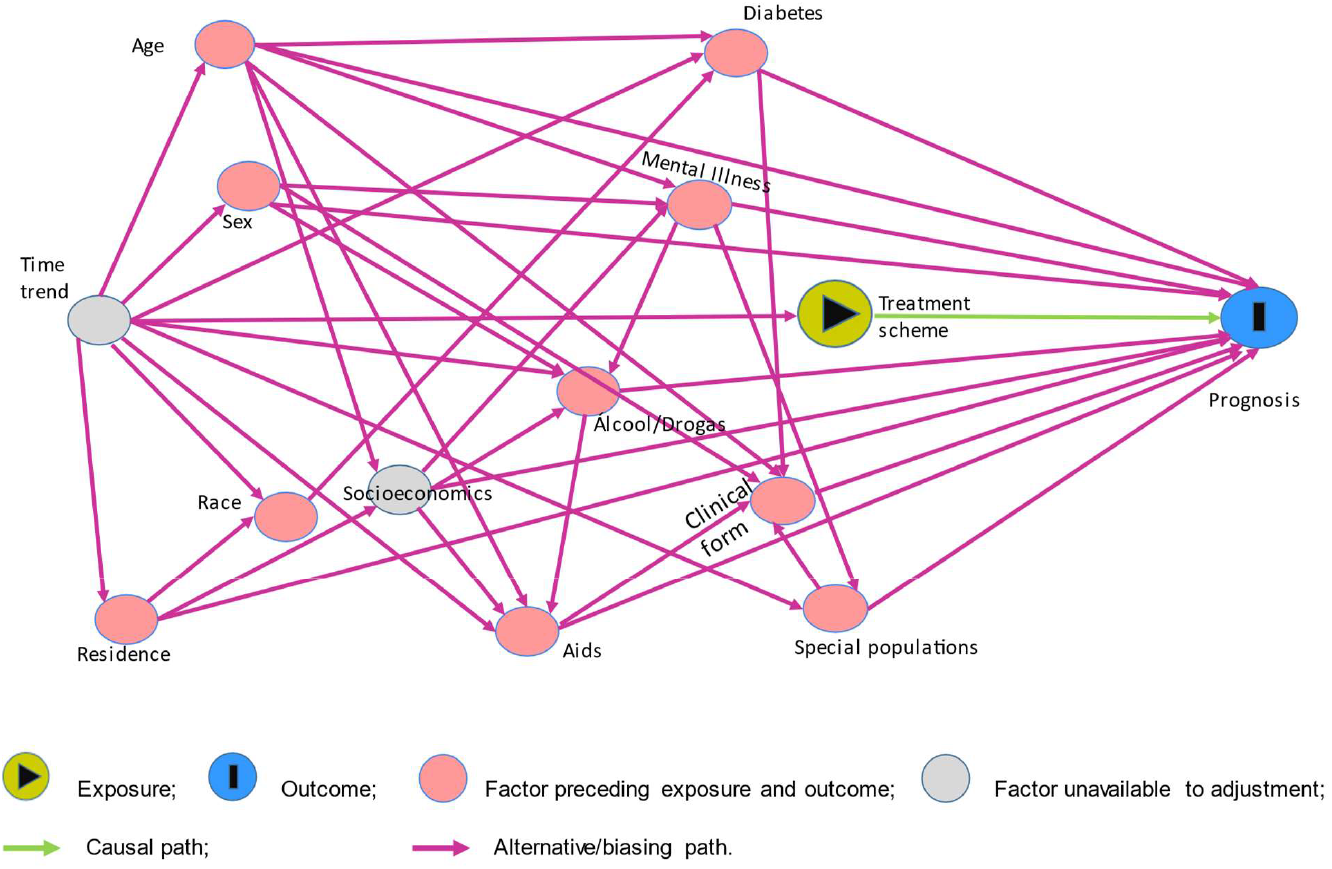
Directed acyclic graph (DAG) of causal pathways relevant to estimate the effect of treatment regimen on tuberculosis prognosis in Brazil.

**Figure 2.**
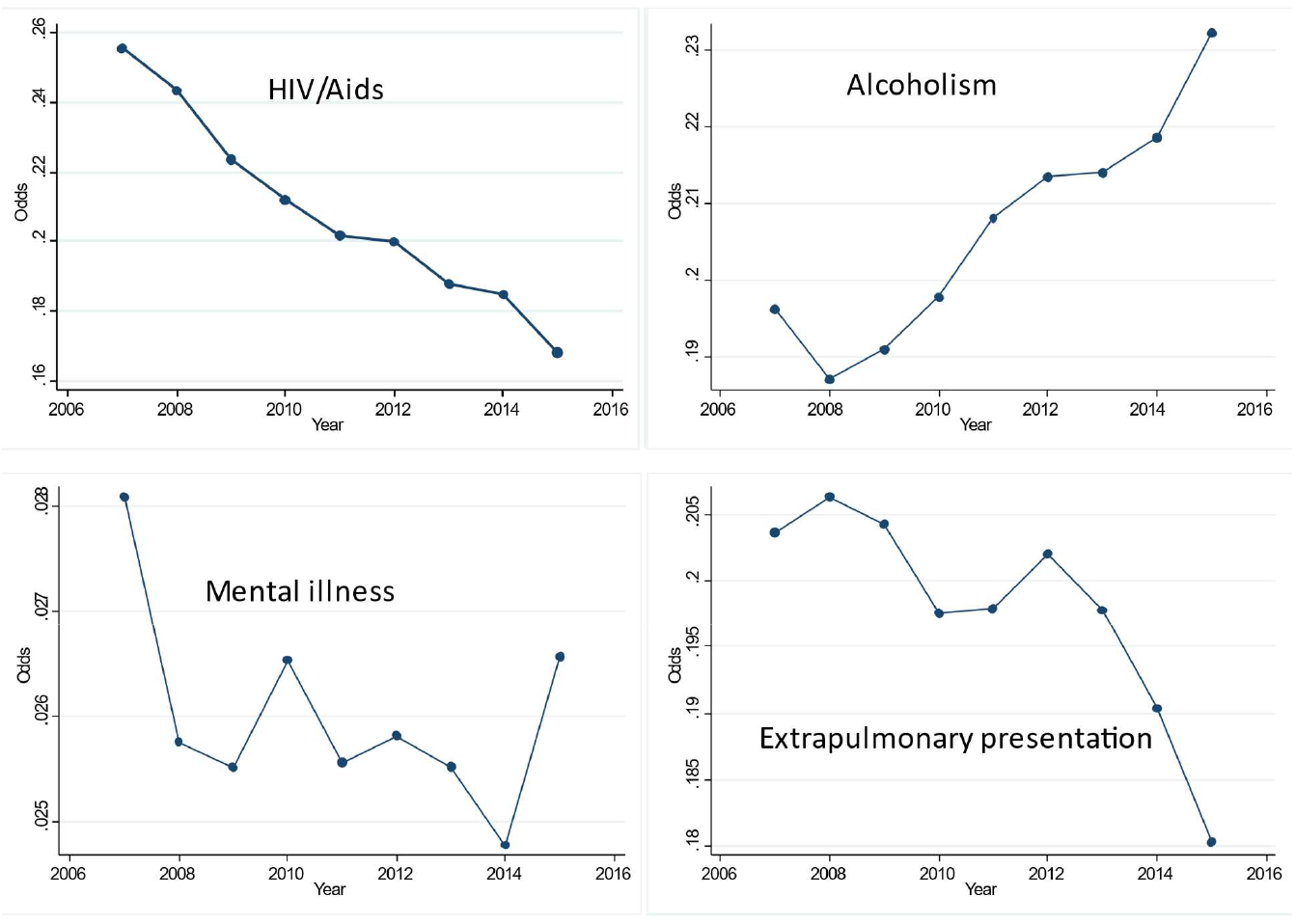
Trends of some covariates in patients with tuberculosis who started treatment in the 2007 to 2015 period.

Therefore, in addition to adjusting for the covariates indicated by the DAG, we decided to restrict the years of the study to the minimum required to obtain a sample with high power, maintaining a reduced temporal distance between the treatment groups. This was intended to minimize residual confounding.

Based on the results of a preliminary analysis, a sample of 148,407 patients was calculated in each of the treatment regimens to document an absolute difference in mortality of 0.35% (taking 9.7% mortality as a reference), with a confidence level of 95% and a power of 90%. Consequently, we chose to include only patients who started treatment between 2008 and 2013, resulting in 145,528 for the reference and 161,264 for the current regimens.

### Imputations

In the database, patients with missing data, in any of the covariates suggested for adjustment, exhibited 15% greater odds of dying (OR: 1.15; 95% CI: 1.11 – 1.19), when adjusted for covariates (with complete data) of treatment (current vs baseline regimen), age, incarcerated population, disease form, and conditioning by state in a multilevel model. Therefore, the imputation of data for the estimates of interest was considered justified.^18^

For estimates that did not require imputation of the outcome (such as mortality), multivariate imputation of covariates was performed with chained equations using logistic regressions for the variables of sex, alcoholism, mental illness, and diabetes; and multinomial regression for a variable of three categories of HIV/AIDS. A three-category variable for cure vs. lost to follow-up vs. other outcomes was also included, leaving transfers and unknown outcome (MOD) as the same kind of missing data, which was imputed with multinomial regression, to contribute to the prediction of missing data for the other variables. Variables with complete data but included as predictors of missing data were their year of diagnosis, age, being an inmate, clinical form (three categories), state, and the all-cause mortality outcome.

For the analysis of secondary outcomes that contained missing data, specific imputations were performed following a similar procedure, but including the outcome variable as dichotomous (using the logistic function in the corresponding equation). Mortality was included as independent for the imputation of cure and lost to follow-up (but not for the first composite outcome because it was one of its components).

### Effect estimations

After defining the most functional forms, all covariates suggested by the final DAG were included in a regression model to estimate the association between the treatment regimen and each of the outcomes. For these purposes, estimation options were used with imputed data in multilevel logistic models of mixed effects, with the state as a cluster.

In each model, the measure of association obtained was the Odds Ratio (OR). The relative risk (RR) was calculated using the following formula:^19^

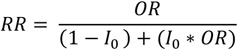

Where *I*_0_ represents the incidence of the corresponding outcome in the reference treatment group.

Furthermore, we evaluated the association between the treatment regimen and mortality outcome using a procedure based on a propensity score.^20^ This score corresponded to the value predicted by a multilevel logistic regression model whose dependent variable was the treatment regimen. In this model, the states were the aggregation units, and the independent variables were the individual risk factors suggested by the DAG. Based on this score, estimates of the effect of the treatment regimen on the outcomes were obtained through analysis stratified by the vigintiles of the score.

We prespecified that the study conclusions would focus on assessing the primary outcome. However, the analysis of secondary outcomes was intended to understand the phenomena involved in the main effect. As eleven estimates were made for secondary outcomes, and taking a value of 0.05 as a reference, we decided to set the significance level at 0.005 (≈0.05/11).

### Effect by subgroups

Given that the current regimen is easier to comply with, it could have a greater effect on groups that have difficulties adhering to it or whose prognosis differs from the rest of the population. Therefore, we evaluated the heterogeneity of treatment effects according to the variables of age, sex, prison population, and HIV/AIDS (which was evaluated as a variable of two and three categories). Regression models adjusted for the variables suggested in the DAG were specified for these purposes, adding the corresponding interaction terms. This analysis was applied exclusively to the all-cause mortality outcome.

For each statistic of the interaction terms, the significance level was fixed *a priori* following the formula of 0.05/k, where k refers to the number of tests. Consequently, a significance level of 0.01 was preset to consider an effect modification.

Technicians from the Ministry of Health linked the databases as part of the surveillance activities of the National Tuberculosis Control Program. The data accessed by the researchers can be accessed upon formal request to the Ministry of Health via “Plataforma Fala.br” and did not contain patient identification. In these cases, Brazilian legislation exempts submission to an ethics committee. All procedures were conducted in accordance with Resolutions 466/12 and 510/2016 of the National Health Council.

## RESULTS

The analyzed sample included 306,792 patients (Figure 3). Compared to the reference group, during the current treatment period, significantly less missing data (categories referred to as unknown or missing) was observed of covariates necessary for adjustment including alcoholism, diabetes, mental illness, and especially HIV/AIDS, dropping from 44.5% to 26.7% (Table 1).

**Figure 3.**
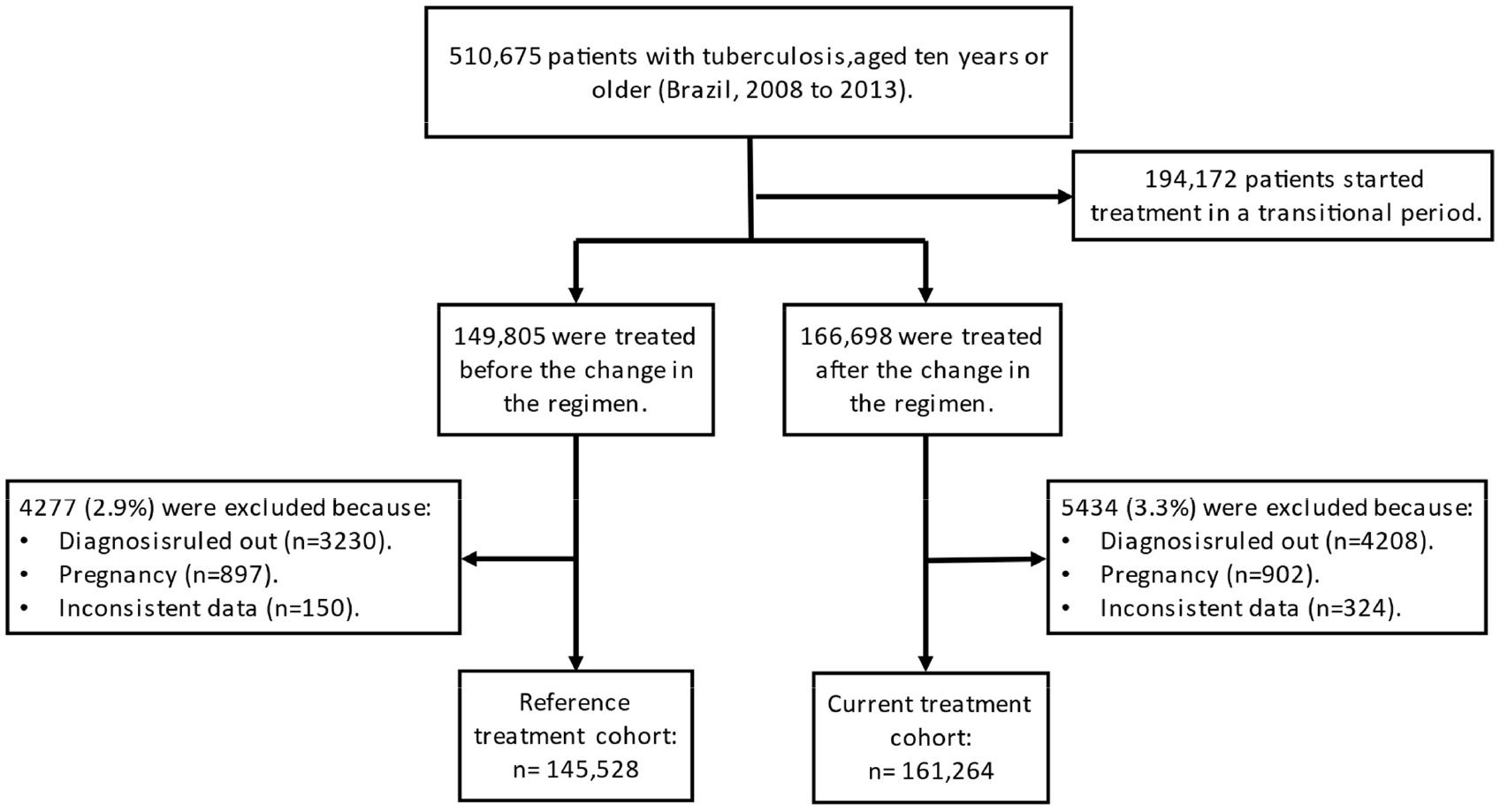
flowchart of patient selection and exclusions.

**Table 1.**
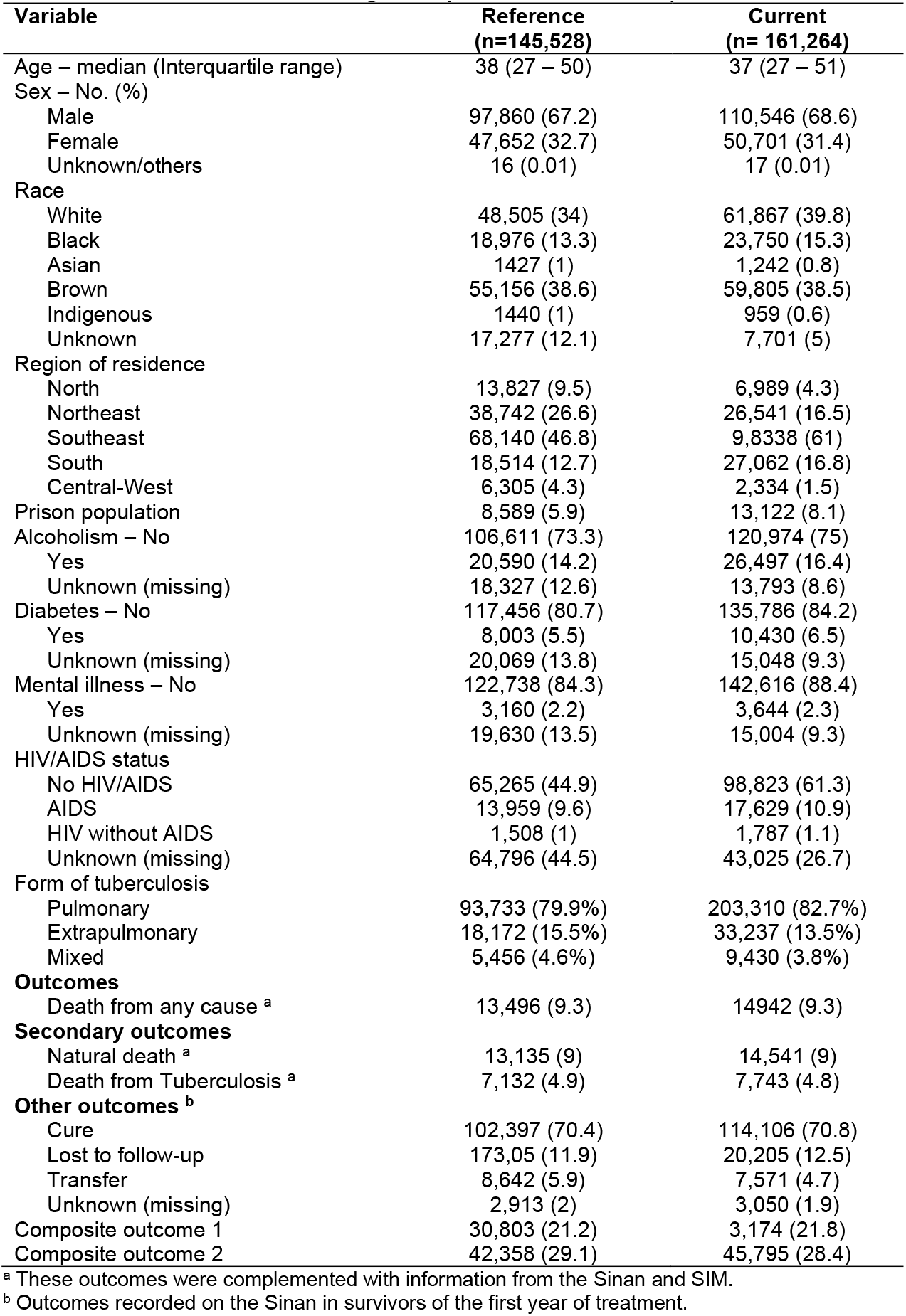
Description of patients with tuberculosis according to the treatment regimen (Brazil, 2008 – 2013).

In the multilevel model (adjusted for the variables suggested by the DAG), compared to the reference group, the current treatment was not associated with significant changes in all-cause mortality (RR: 1.01; 95%CI: 0.98 – 1.04), natural death (RR: 1.01; 95%CI: 0.98 – 1.04), or death from tuberculosis (RR: 0.98; 95%CI: 0.95 – 1.02). The results were similar with the analysis stratified by the 20 quantiles of the propensity score, with RRs of 1.01 (95%CI: 0.99 – 1.04), 1.01 (95%CI: 0.99 – 1.04), and 0.99 (95%CI: 0.96 – 1.02), respectively (Table 2).

**Table 2.** Association between tuberculosis treatment regimen (current/reference) and clinical outcomes.

| Outcome | RR | p-value |
| --- | --- | --- |
| **Primary** |  |  |
| Death from any cause | 1.01 (0.98 – 1.04) | 0.79 |
| **Secondary** |  |  |
| Natural death | 1.01 (0.98 – 1.04) | 0.43 |
| Death from tuberculosis | 0.98 (0.95 – 1.02) | 0.31 |
| Cure<sup>a</sup> | 1 (0.99 – 1) | 0.51 |
| Cure<sup>b</sup> | 0.99 (0.98 – 0.99) | <0.001 |
| Cure<sup>c</sup> | 0.98 (0.98 – 0.99) | <0.001 |
| Lost to follow-up<sup>a</sup> | 1.06 (1.04 – 1.09) | <0.001 |
| Lost to follow-up<sup>b</sup> | 0.99 (0.98 – 1.01) | 0.48 |
| Lost to follow-up<sup>c</sup> | 1.06 (1.03 – 1.08) | <0.001 |
| Composite outcome 1<sup>a</sup> | 1.04 (1.02 – 1.06) | <0.001 |
| Composite outcome 1<sup>c</sup> | 1.03 (1.02 – 1.05) | <0.001 |
| Composite outcome 2<sup>d</sup> | 1 (0.99 – 1.01) | 0.71 |

<sup>a</sup> Considering transfer and missing outcome data (MOD) as the absence of this outcome.
<sup>b</sup> Considering transfer and MOD as the occurrence of this outcome.
<sup>c</sup> Imputing transfers and MOD.
<sup>d</sup> Considering MOD as the occurrence of outcome and transfer as the absence of this outcome.

Furthermore, the use of a dichotomous variable to represent HIV/AIDS, rather than a 3-category variable, led to a similar estimates of the current treatment effect (e.g., RR all-cause mortality: 1.01; 95%CI: 0.98 – 1.04; p=0.32).

During the current regimen, the frequency of the outcome defined as transfer or unknown reduced from 8.5% to 7.1%, i.e., a relative reduction of 17.2% (95% CI: 15.1% – 19.2%). These losses of information about the final situation in the Sinan could potentially explain the associations between treatment and some secondary outcomes. Specifically, in relation to cure, when transfers and MOD were considered as the absence of this event, there were no differences between treatments. However, when they were considered as a cure or imputed, the outcome was less frequent in the current treatment.

When transfers and MOD (in the Sinan form) were considered, the absence of loss to follow-up or this outcome imputed, lost to follow-up was more frequent in the current treatment. However, when those categories were interpreted as lost to follow-up, there were no differences between treatments. Something similar happened with the unsuccessful composite outcomes that were similar between groups when missing data were considered as the occurrence of the event. When they were considered as absence of the same or imputed, there was a higher frequency with the current treatment (Table 2).

The effect of treatment on mortality did not present a statistically significant heterogeneity attributable to age, sex, prison population, or HIV/AIDS status (Table 3).

**Table 3.** Assessment of the heterogeneity of the effects of the treatment regimen effects on all-cause mortality.

| Suggested effect modifier | Ratio of ORs | p-value |
| --- | --- | --- |
| Age | 1.01 (0.99 – 1.03) | 0.31 |
| Sex male | 1.04 (0.98 – 1.1) | 0.23 |
| Prison population | 0.91 (0.79 – 1.04) | 0.16 |
| HIV/Aids | 0.92 (0.86 – 0.99) | 0.03 |

## DISCUSSION

Changes in the treatment regimen, including ethambutol and simplification of drug presentation using combination fixed-dose, were not associated with all-cause mortality in patients with tuberculosis. These findings were consistent when evaluating secondary mortality outcomes, such as natural death and death from tuberculosis.

From a methodological perspective, the present work emphasizes the importance of evaluating population trends using individual data. Our results suggest that the previously reported increase in mortality should not be attributable to the new treatment regimen.^8^ Instead, that trend can be explained by changes in the determinants of prognosis, some of which were observed in the present study to be strongly affected by time. Therefore, it was essential to restrict the observation period to minimize residual confounding either by errors in measuring covariates or by unknown or unmeasured causal pathways.

Additionally, the imputation of covariates avoided selection bias due to missing information. A complete-case analysis suggested a modest reduction in mortality with the new treatment regimen (OR=0.96; 95%CI: 0.93 – 0.98 [analysis not shown]). However, subjects with missing data were less frequent in the current regimen and had a different prognosis than those in the reference treatment, regardless of other covariates. Thus, there was a clear indication to prefer an analysis with imputed data.^18^ Therefore, the present analysis sought to mitigate the impact of missingness that was especially frequent for the HIV/AIDS variable, which was essential for adjustment.

When analyzing other outcomes, the current treatment period was associated with fewer cures and more lost to follow-up, as recorded in the Sinan. However, the sensitivity analysis indicates that the reduction of transfers and unknown outcomes could explain these associations during the second period. By the way, we noticed the positive trend of information qualification, with a decrease in missing data as an indicator of improvements in the disease surveillance systems in the country.

## Limitations

The retrospective design of this study may pose challenges related to unmeasured variables or changes in recording patterns of relevant factors. For example, the incorporation of a new diagnostic method at the end of the study period may be associated with increased identification of cases of drug-resistant tuberculosis during the current treatment period. These resistant cases require a different treatment regimen than those compared. However, we did not exclude from the analysis cases classified in the Sinan as drug-resistant tuberculosis, as this could differentially affect the periods under study. To mitigate the effect of this limitation, the analyzes were restricted to new cases, since all of them should have started the basic treatment regimen.

The current study did not aim to evaluate the effect of the new regimen on the incidence of drug resistance in tuberculosis, which was one of the objectives of changing the regimen. The analysis made it possible to assess the safety of its implementation among new cases and to compare the mortality of people undergoing treatment. Thus, the probabilistic relationship of large databases was a strategy for qualifying information on deaths.

The frequency of drug resistance was expected to increase in the second period and these patients may have a worse prognosis. We believe that if conditioning with this variable had been possible, the current treatment regimen would be associated with a better prognosis. Therefore, considering the probable emergence of resistance (difficult to measure accurately), we may interpret that the change in the treatment regimen could have prevented the increase in mortality.

On the other hand, transfers and the difficulty of accurately dating the time of loss to follow-up make the evaluated secondary outcomes susceptible to classification errors. For these reasons, it is important that the main conclusions are based on all-cause mortality.

### Implications of the study

This study evaluated the effect of changing the tuberculosis treatment regimen in the real settings of routine health services in the country. Based on individual data, this work raised the level of evidence, controlling or mitigating confounding phenomena. Additionally, the study was carried out using large databases, with high power to identify associations and obtain precise estimates. Finally, our analysis is expected to serve as a methodological reference on the use of surveillance information systems to evaluate public health interventions and programs in Brazil.

## CONCLUSION

In the study population, the changes made in the tuberculosis treatment regimen were not associated with an increase in mortality. Differences in the frequency of loss to follow-up and cure can be explained by an improvement in disease surveillance in the country, which qualified the information on the treatment outcome.

## Data Availability

All data produced in the present study are available upon reasonable request to the authors.

## AUTHORS’ CONTRIBUTION

FADQ participated in the conception and design of the study, performed data analysis, and prepared the first version of the manuscript. PB and RSP made substantial contributions to data acquisition. PB, KBA, DMP, DAS, FDC, and RSP participated in the design, discussed the results of the first version, and revised the manuscript. All authors provided relevant information for writing, performed revisions, and read and approved the final manuscript. Therefore, each author participated sufficiently in the work to take public responsibility for appropriate portions of the content and, thus, agreed to be accountable for all aspects in ensuring that questions related to the accuracy or integrity of any part of the work are appropriately investigated and resolved.

## ACKNOWLEDGMENTS

The authors thank Rodrigo Couto and Daiane Alves for their suggestions. FADQ also thanks his daughter, Ana Maria, for her help in drawing the DAG.

